# Increased Risk of Ischemic Stroke in Amyotrophic Lateral Sclerosis: A Nationwide Cohort Study in South Korea

**DOI:** 10.1101/2023.08.11.23294006

**Authors:** Soonwook Kwon, Bongseong Kim, Kyung-Do Han, Wonyoung Jung, Eun Bin Cho, Dong Wook Shin, Ju-Hong Min

## Abstract

**Background:** We investigated the risk of ischemic stroke in ALS and analyzed the effect of ALS-related physical disability using the Korean National Health Insurance Service database.

**Methods:** A total of 2,252 ALS patients diagnosed between January 1, 2012, and December 31, 2015, and 1:10 age- and sex-matched control populations were included. After selection of cases that participated in the national health check-up programs, 659 ALS patients and 10,927 non-ALS participants were remained. Newly developing ischemic stroke as primary outcome was also defined by the ICD code and the incidence probability was assessed using by the Kaplan– Meier method during the follow-up duration. A Cox hazard regression model was used to examine the hazard ratios (HRs) for ischemic stroke in ALS after adjusting for potential confounders.

**Results:** ALS patients were younger than the control group (60.3 ± 10.2 years vs. 61.4 ± 10.5 years, p = 0.008), and the proportion of male patients was similar between the two groups (61.0% vs. 62.5%, p = 0.447). ALS patients were more likely to have a lower body mass index (23.1 ± 2.92 vs. 24.0 ± 3.00, p < 0.001) and be non-drinkers (64.2% vs. 57.4%, p = 0.002) than the controls. In ALS patients, the incidence of ischemic stroke was 7.8 per 1,000 person-years, and the adjusted HR of ischemic stroke was 3.67 (95% confidence interval 2.02−6.67) compared with the control group. The risk of ischemic stroke did not differ by the presence of disability in ALS patients.

**Conclusions:** Our findings suggest that ALS patients face increased risk of ischemic stroke compared with controls, but the risk did not differ by the presence of disability in ALS.

## Introduction

Amyotrophic lateral sclerosis (ALS) is a representative neurodegenerative disease that affects both upper and lower motor neurons. The mechanism of ALS is not fully understood, but the mislocalization and aggregation of transactive response DNA-binding protein ∼43 kDa (TDP-43) in the cytoplasm play an important role.^1^ Recently, TDP-43 proteinopathy has been reported to be associated with brain arteriolosclerosis, although the association with TDP-43 and ischemic stroke is unclear.^2^ In addition, increased TDP-43 can exacerbate atherosclerosis by promoting inflammation and lipid uptake of macrophages, which can increase the risk of stroke.^3^

Previously, the prevalence of stroke in ALS patients has been reported to be 1.6–8.0% in case-control studies, with inconsistent results (Supplementary Table 1).^4, 5^ In a study of 500 patients with motor neuron disease (MND) (ALS, 76.5%) in Portugal, the prevalence of stroke (both hemorrhagic and ischemic stroke) did not differ from that of the controls, regardless of the region of onset.^4^ However, another study of 200 patients with MND (ALS, 92.4%) in Germany suggested that the prevalence of ischemic stroke was higher in MND groups compared with controls.^5^ On the other hand, there has been a report that prior ischemic stroke increased the risk of ALS; in a cohort study in England, the relative risk of ALS was found to be 1.31 times higher than the expected number.^6^ This study suggests an association between ischemic stroke and ALS. However, the risk of ischemic stroke after the onset of ALS has not yet been reported.

In the present study, we investigated the risk of developing ischemic stroke in ALS patients compared with control populations and the effect of ALS-related physical disability on the risk using the Korean National Health Insurance Service (NHIS) database.

## Methods

### Data sourcing and study setting

The NHIS is a single-payer program that covers approximately 97% of the entire Korean population; the remaining 3% are covered as Medicaid beneficiaries.^7^ The NHIS database contains health information on demographics, health care use, and claims data, including diagnosis codes defined by the International Classification of Diseases-Tenth Revision (ICD-10), and links to a death registry database to manage the qualification of enrollees.^8^ In addition, the NHIS provides biennial national health screenings for all beneficiaries aged 40 and older that include: a health questionnaire for lifestyle factors (exercise, alcohol consumption, and smoking), anthropometric measurements (blood pressure, body weight, and height), and laboratory tests (blood glucose, lipid profile, serum creatinine, and other health indicators).^9^

### Study participants

Individuals who were first diagnosed with ALS between January 1, 2012, and December 31, 2015, were enrolled in the present study. An operational definition of ALS was designed based on both ICD-10 diagnosis codes and information in the Rare Intractable Disease (RID) management program in South Korea. The RID program provides a copayment reduction of up to 10% for various rare and intractable diseases including ALS.^10^ To be registered in the RID program, a diagnosis certification submitted by a neurologist is required based on the Awaji criteria or revised El Escorial criteria. A total of 2,252 ALS patients were identified according to the following criteria (Figure 1): (i) ≥2 outpatient or hospitalization claims with the ICD-10 code G12.21 for ALS and (ii) registration in the RID with code V123 for ALS. To obtain variables such as lifestyle factors, anthropometric measurements, and laboratory results, patients who participated in the national health check-up programs within two years from the index date were finally included. The index date of ALS cases was designated as the first date of claims for the ALS diagnosis with both the ICD and RID codes. Among these patients, those who (i) were younger than 20 years (N=11), (ii) had a previous history of ischemic stroke or myocardial infarction before an ALS diagnosis (N=484), or (iii) had unavailable data for variables (N=29) were excluded. Finally, 659 ALS patients were included (Figure 1).

**Figure 1.**
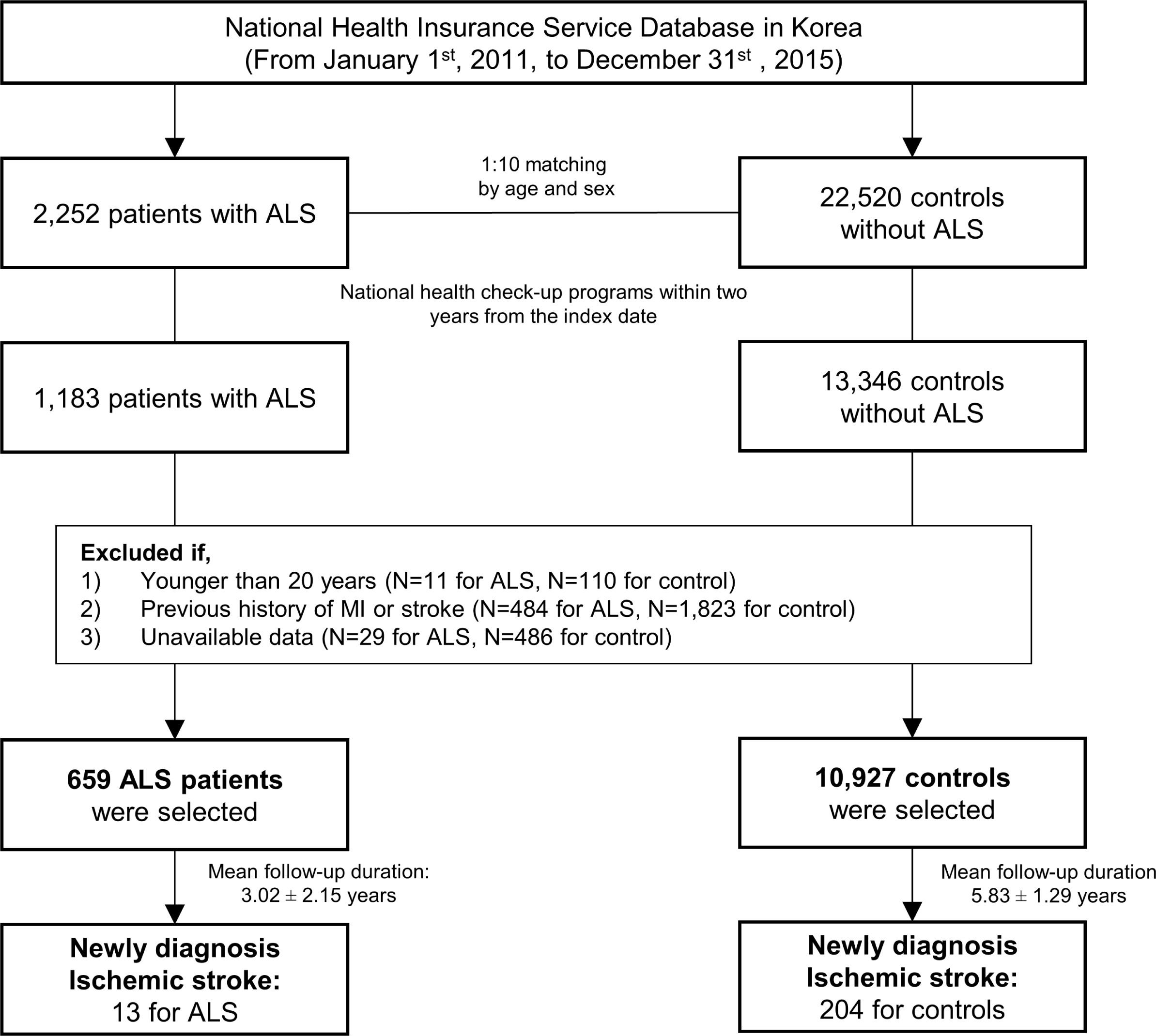
Flow diagram of study population enrollment.

For the control group, we obtained an initial control pool of 22,520 subjects who did not have an ALS diagnosis matched by age, sex, and index year (1:10 matching). In the control group, 13,346 participants participated in the national health check-up programs within two years from the index date. After applying identical exclusion criteria, 10,927 non-ALS participants were included in the analysis.

This study was approved by the Institutional Review Board of Samsung Medical Center (File No. SMC-2020-12-108). The review board waived the requirement for written informed consent because publicly open and anonymous data were used for the analysis and retrospective features.

### Assessment of Disability

The severity of ALS was determined by the disability grade registered in the National Disability Registration System (NDRS) of the Ministry of Health and Welfare.^11^ Because the NDRS provides social welfare benefits based on predefined criteria for disability and an objective medical assessment using a disability grading system, most ALS patients apply to be registered with the NDRS. The NDRS includes information on the type and severity of disability, which are legally defined in Korea. The types of disability are broadly divided into physical disabilities and mental disabilities, with a total of 15 legally defined types in detail. Physical disabilities include disabilities of the extremities, vision, hearing, and speech and language, and disabilities due to brain injury, facial deformity, renal failure, heart problems, liver disease, respiratory problems, ostomy, and epilepsy. Mental disabilities include intellectual disorders, autism, and mental disorders.^11^ The degree of disability is classified from grade 1, the most severe, to grade 6, the least severe.

ALS is classified as a disability of the extremities or/and brain injury among physical disabilities, and the disability severity is assessed by two or more specialists in neurology, neurosurgery, or rehabilitation medicine based on the submitted documents (Supplementary Tables 2 and 3). In this study, the disabled ALS group was defined as ALS patients who were registered in the NDRS within two years from the index date. The non-disabled ALS group was defined as ALS patients who had not yet developed disabilities meeting the criteria of NDRS.^11^

### Primary outcome

The primary outcome of this study was newly diagnosed ischemic stroke, defined as at least one hospitalization for ICD-10 codes I63 or I64 and claims for brain imaging including brain computed tomography or magnetic resonance imaging during the follow-up duration.^12^ The cohort was followed from the index date to the date of ischemic stroke onset, death, tracheostomy,^13^ or the end of the study (December 31, 2019), whichever came first.

### Covariates

Hypertension, diabetes mellitus (DM), and dyslipidemia were defined based on claims, prescription information, and data from biennial national health screenings prior to the index date. The Charlson Comorbidity Index (CCI) was also calculated based on ICD-10 codes.^14^ Lifestyle factors (alcohol consumption, smoking, and regular exercise), anthropometric measurements (height, weight, and body mass index [BMI]), and socioeconomic information (income and region) were obtained by linkage to the results of national health check-up programs within two years before the index date.

### Statistical analysis

Descriptive statistics were calculated as the mean ± standard deviation (SD) for continuous variables and the number (percentage) for categorical variables. We conducted Student’s t-test and one-way analysis of variance for continuous variables and χ2 test for categorical variables when comparing two groups.

The probability of ischemic stroke incidence was assessed using the Kaplan–Meier method. The association between ALS and ischemic stroke incidence was estimated using Cox proportional hazards regression models adjusted for age, sex, income, CCI, region, smoking, alcohol consumption, regular exercise, and comorbidities (hypertension, DM, and dyslipidemia). Adjusted hazard ratios (aHRs) with 95% confidence intervals (CI) were calculated.

Subgroup analyses were performed according to sex, age (< 65 and ≥ 65 years), and BMI (<25 and ≥25). Multiplicative interactions between the stratifying variables were also tested. All statistical analyses were conducted using SAS statistical package version 9.4 (SAS Institute Inc., Cary, NC, USA), and a p-value < 0.05 was considered statistically significant.

## Results

Table 1 shows the results of basic characteristics of the ALS and control groups. The mean age of the ALS group was younger than that of the control group (60.3 ± 10.2 years vs. 61.4 ± 10.5 years, p = 0.008). The mean BMI was lower in the ALS group (23.1 ± 2.9 vs. 24.0 ± 3.0, p < 0.001) and the presence of hypertension, DM, and dyslipidemia did not differ between the ALS and control groups. CCI was higher in the ALS group (3.7 ± 2.4 vs. 1.5 ± 1.7, p < 0.001). Smoking status did not differ between the two groups (p = 0.271), but the proportion of non-drinkers was higher in the ALS group than in the control group (62.4% vs. 57.4%, p = 0.002). The proportion of patients with low income was higher in the ALS group than in the control group (22.8% vs. 19.3%, p = 0.029). There were no significant differences in baseline characteristics between the disabled and non-disabled ALS groups. The mean durations of follow-up from the index year were 3.0 ± 2.2 years and 5.8 ± 1.3 years in the ALS and control groups, respectively (Figure 1).

**Table 1.**
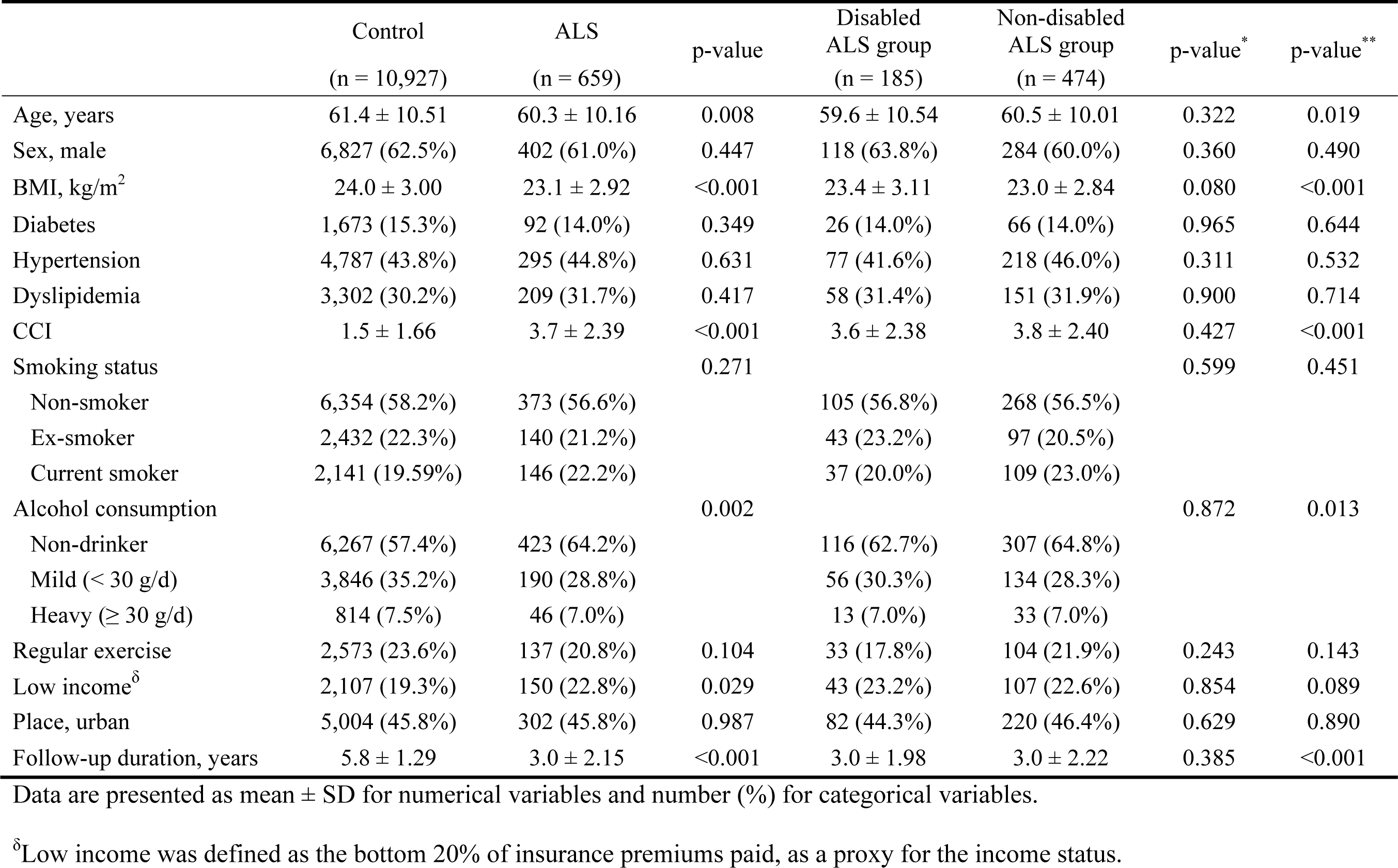
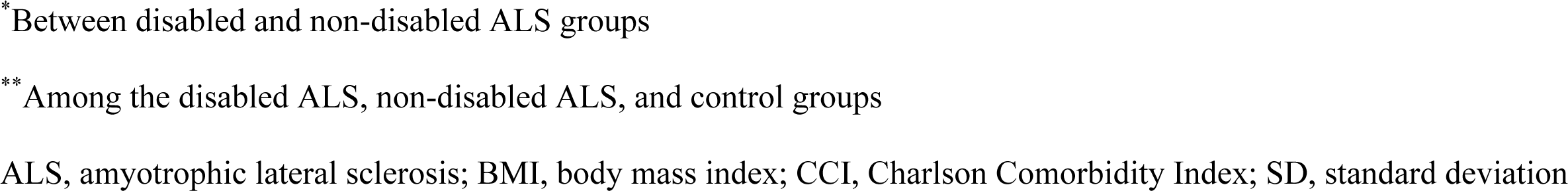
Demographics of ALS patients and the control population.

|  | Control<br>(n = 10,927) | ALS<br>(n = 659) | p-value | Disabled<br>ALS group<br>(n = 185) | Non-disabled<br>ALS group<br>(n = 474) | p-value* | p-value** |
| --- | --- | --- | --- | --- | --- | --- | --- |
| Age, years | 61.4 ± 10.51 | 60.3 ± 10.16 | 0.008 | 59.6 ± 10.54 | 60.5 ± 10.01 | 0.322 | 0.019 |
| Sex, male | 6,827 (62.5%) | 402 (61.0%) | 0.447 | 118 (63.8%) | 284 (60.0%) | 0.360 | 0.490 |
| BMI, kg/m<sup>2</sup> | 24.0 ± 3.00 | 23.1 ± 2.92 | <0.001 | 23.4 ± 3.11 | 23.0 ± 2.84 | 0.080 | <0.001 |
| Diabetes | 1,673 (15.3%) | 92 (14.0%) | 0.349 | 26 (14.0%) | 66 (14.0%) | 0.965 | 0.644 |
| Hypertension | 4,787 (43.8%) | 295 (44.8%) | 0.631 | 77 (41.6%) | 218 (46.0%) | 0.311 | 0.532 |
| Dyslipidemia | 3,302 (30.2%) | 209 (31.7%) | 0.417 | 58 (31.4%) | 151 (31.9%) | 0.900 | 0.714 |
| CCI | 1.5 ± 1.66 | 3.7 ± 2.39 | <0.001 | 3.6 ± 2.38 | 3.8 ± 2.40 | 0.427 | <0.001 |
| Smoking status |  |  | 0.271 |  |  | 0.599 | 0.451 |
| Non-smoker | 6,354 (58.2%) | 373 (56.6%) |  | 105 (56.8%) | 268 (56.5%) |  |  |
| Ex-smoker | 2,432 (22.3%) | 140 (21.2%) |  | 43 (23.2%) | 97 (20.5%) |  |  |
| Current smoker | 2,141 (19.59%) | 146 (22.2%) |  | 37 (20.0%) | 109 (23.0%) |  |  |
| Alcohol consumption |  |  | 0.002 |  |  | 0.872 | 0.013 |
| Non-drinker | 6,267 (57.4%) | 423 (64.2%) |  | 116 (62.7%) | 307 (64.8%) |  |  |
| Mild (< 30 g/d) | 3,846 (35.2%) | 190 (28.8%) |  | 56 (30.3%) | 134 (28.3%) |  |  |
| Heavy (≥ 30 g/d) | 814 (7.5%) | 46 (7.0%) |  | 13 (7.0%) | 33 (7.0%) |  |  |
| Regular exercise | 2,573 (23.6%) | 137 (20.8%) | 0.104 | 33 (17.8%) | 104 (21.9%) | 0.243 | 0.143 |
| Low income<sup>δ</sup> | 2,107 (19.3%) | 150 (22.8%) | 0.029 | 43 (23.2%) | 107 (22.6%) | 0.854 | 0.089 |
| Place, urban | 5,004 (45.8%) | 302 (45.8%) | 0.987 | 82 (44.3%) | 220 (46.4%) | 0.629 | 0.890 |
| Follow-up duration, years | 5.8 ± 1.29 | 3.0 ± 2.15 | <0.001 | 3.0 ± 1.98 | 3.0 ± 2.22 | 0.385 | <0.001 |

Data are presented as mean ± SD for numerical variables and number (%) for categorical variables.
<sup>δ</sup>Low income was defined as the bottom 20% of insurance premiums paid, as a proxy for the income status.
\*Between disabled and non-disabled ALS groups
\*\* Among the disabled ALS, non-disabled ALS, and control groups
ALS, amyotrophic lateral sclerosis; BMI, body mass index; CCI, Charlson Comorbidity Index; SD, standard deviation

### Risk of ischemic stroke in the ALS group compared with the control group

During the follow-up period, there were 13 ischemic strokes in the ALS group and 204 in the control group (Table 2). The incidence rates were 7.8/1,000 person-year (PY) in the ALS group and 3.2/1,000 PY in the control group. The unadjusted HR and adjusted HR (aHR) of ischemic stroke were 2.5 (95% CI 1.44−4.47) and 3.7 (95% CI 2.02−6.67), respectively. The aHRs of ischemic stroke were similar in both the disabled and non-disabled ALS groups. Kaplan–Meier analysis showed that the incidence probabilities of ischemic stroke in the ALS group were higher than those in the control group (Figure 2A). Between the disabled and non-disabled ALS groups, the incidence probabilities of ischemic stroke did not differ (Figure 2B).

**Table 2.** Incidence rates and risk of ischemic stroke in ALS patients.

|  | N | Ischemic stroke | Person-year | Incidence rate<sup>δ</sup> | Unadjusted HR<br>(95% CI) | Adjusted HR<sup>†</sup><br>(95% CI) |
| --- | --- | --- | --- | --- | --- | --- |
| ***Comparison between the ALS and control groups*** |  |  |  |  |  |  |
| Control | 10,927 | 204 | 64,111.0 | 3.2 | 1 (ref.) | 1 (ref.) |
| ALS group | 659 | 13 | 1,671.6 | 7.8 | 2.54 (1.44, 4.47) | 3.67 (2.02, 6.67) |
| ***Comparison between the disabled ALS, non-disabled ALS, and control groups*** |  |  |  |  |  |  |
| Control | 10,927 | 204 | 64,111.0 | 3.2 | 1 (ref.) | 1 (ref.) |
| Non-disabled ALS group | 474 | 9 | 1,206.4 | 7.5 | 2.43 (1.24, 4.76) | 3.67 (1.82, 7.40) |
| Disabled ALS group | 185 | 4 | 465.2 | 8.6 | 2.81 (1.04, 7.59) | 3.68 (1.34, 10.08) |

<sup>δ</sup>Incidence rates of 1,000 person-years
<sup>†</sup>Adjusted for age, sex, Charlson Comorbidity Index, place, income, diabetes, hypertension, dyslipidemia, smoking, alcohol consumption, and regular exercise
ALS, amyotrophic lateral sclerosis; CI, confidence interval; HR, hazard ratio

**Figure 2.**
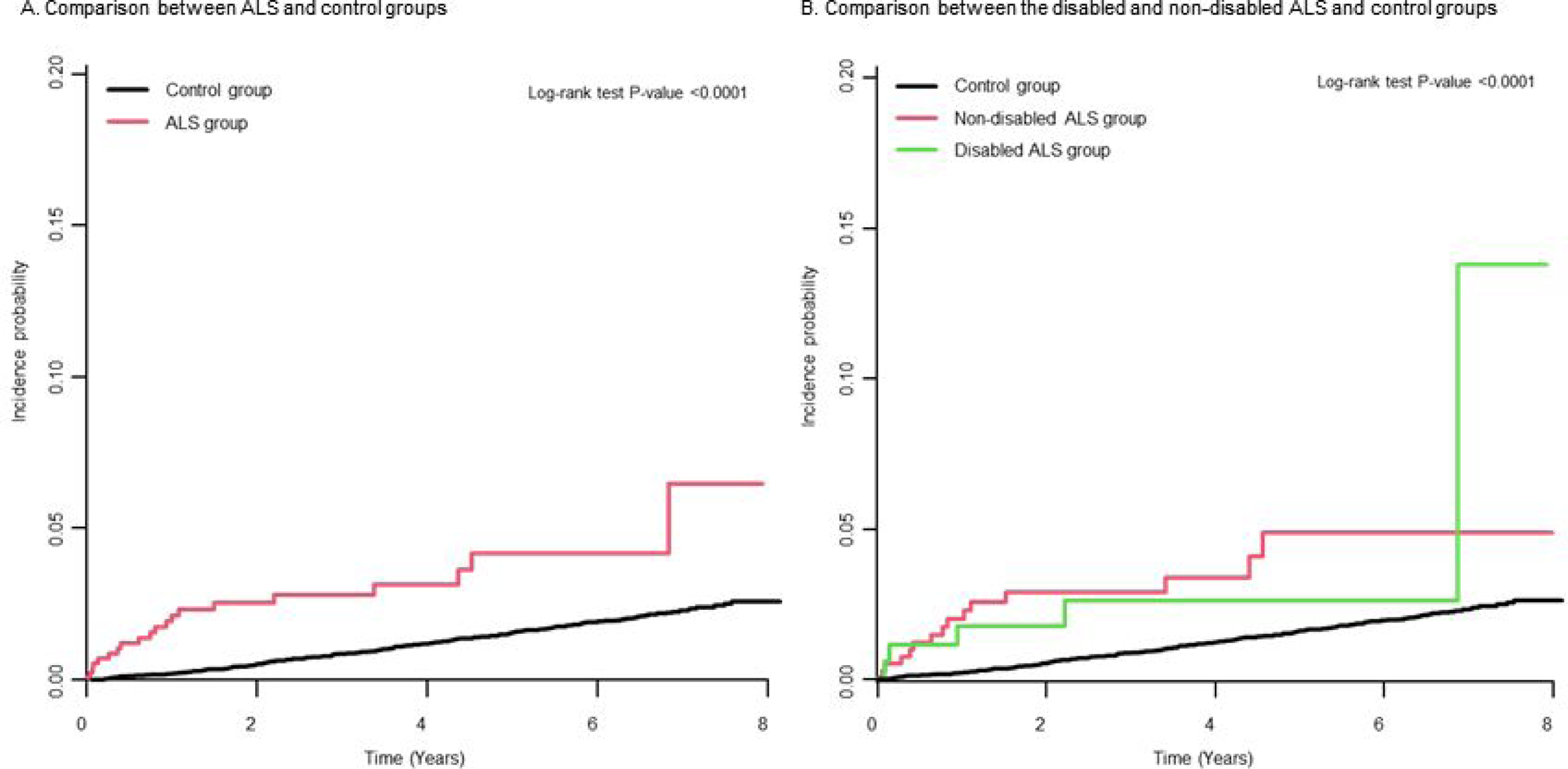
Kaplan–Meier curves of the probability of the incidence of ischemic stroke in ALS patients and a control group. (A) Compared with the control group, ALS patients showed a higher incidence probability of ischemic stroke during the follow-up period. (B) The ALS group from (A) was divided into two groups: a disabled ALS group and a non-disabled ALS group.

### Subgroup analyses

In the stratified analyses, there was no difference in risk according to age, sex, or BMI. An increased risk of ischemic stroke was consistently observed in each subgroup, but that was not shown in ALS patients with a BMI ≥ 25 (Table 3).

**Table 3.** Subgroup analysis of ischemic stroke in ALS patients stratified by age, sex, and BMI.

|  |  | N | Ischemic stroke | Duration | Incidence rate<sup>δ</sup> | Adjusted HR<sup>†</sup><br>(95% CI) | p for interaction |
| --- | --- | --- | --- | --- | --- | --- | --- |
| Age < 65 years | Control | 6,375 | 54 | 37,566.0 | 1.4 | 1 (ref.) | 0.792 |
|  | ALS | 421 | 6 | 1,234.8 | 4.9 | 4.00 (1.68, 9.51) |  |
| Age ≥ 65 years | Control | 4,552 | 150 | 26,545.0 | 5.7 | 1 (ref.) |  |
|  | ALS | 238 | 7 | 436.9 | 16.0 | 3.43 (1.57, 7.51) |  |
| Male | Control | 6,827 | 141 | 39,740.8 | 3.5 | 1 (ref.) | 0.141 |
|  | ALS | 402 | 6 | 937.6 | 6.4 | 2.54 (1.09, 5.90) |  |
| Female | Control | 4,100 | 63 | 24,370.2 | 2.6 | 1 (ref.) |  |
|  | ALS | 257 | 7 | 734.0 | 9.5 | 5.94 (2.65, 13.31) |  |
| BMI < 25 | Control | 7,053 | 127 | 41,380.3 | 3.1 | 1 (ref.) | 0.569 |
|  | ALS | 500 | 10 | 1,236.0 | 8.1 | 4.10 (2.08, 8.10) |  |
| BMI ≥ 25 | Control | 3,874 | 77 | 22,730.7 | 3.4 | 1 (ref.) |  |
|  | ALS | 159 | 3 | 435.6 | 6.9 | 2.79 (0.87, 8.98) |  |

<sup>δ</sup>Incidence rates of 1,000 person-years
<sup>†</sup>Adjusted for age, sex, Charlson Comorbidity Index, place, income, diabetes, hypertension, dyslipidemia, smoking, alcohol consumption, and regular exercise
ALS, amyotrophic lateral sclerosis; BMI; body mass index; CI, confidence interval; HR, hazard ratio

## Discussion

In this study, we explored the risk of ischemic stroke in ALS compared with the general population and investigated the effect of disability severity on the risk of ischemic stroke. This population-based cohort study indicated that ALS patients had 3.7-fold higher risk compared with controls after adjusting for age, sex, CCI, place, income, DM, hypertension, dyslipidemia, smoking, alcohol consumption, and regular exercise. The severity of physical disability did not affect the risk of ischemic stroke in ALS patients.

To our knowledge, this is the first study to show an increased risk of ischemic stroke in ALS. This finding is not well explained by vascular risk factors. Vascular risk factors such as hypertension, DM, dyslipidemia, and smoking did not differ between ALS patients and controls, and ALS patients were more likely to have a lower BMI and be non-drinkers. Previous reports have also shown that vascular risk factors are low in ALS patients.^15, 16^ There are several possible explanations for the increased risk of ischemic stroke not explained by vascular risk factors. First, ischemic stroke can be provoked by paradoxical embolism. Venous thromboembolism is common in ALS patients due to leg weakness and reduced mobility, and the risk of deep vein thrombosis is 3.2 times higher than in non-ALS individuals.^17, 18^ This increased risk of thromboembolism in ALS may explain the higher risk of ischemic stroke. Second, increased systemic inflammation in ALS causes the proatherogenic environment that leads to ischemic stroke. Inflammation, which plays a role in the development of ALS, is not limited to the central nervous system; systemic inflammation is also increased.^1, 19^ Increased interleukin-17 is predominantly proatherogenic,^20^ and increased C-reactive protein and monocyte chemoattractant protein-1 are known to be independently associated with the risk of ischemic stroke.^21^ In addition, TDP-43, which plays a role in the pathogenesis of ALS, can exacerbate atherosclerosis by promoting inflammation and lipid uptake of macrophages.^3^ Lastly, endothelial damage and impaired endothelium repair occur in ALS,^22^ and this endothelial dysfunction may be involved in cerebral small-vessel disease and lacunar infarction due to lipohyalinosis.^23^ Further research is needed to understand the mechanism of increased ischemic stroke in ALS.

Compared with controls, ALS patients had more occurrences of ischemic stroke or myocardial infarction before the index year (21.5% in ALS patients vs. 8.1% in controls), which may suggest that vascular events were already occurring before ALS diagnosis. This is supported by the fact that the pathogenesis of ALS is already advanced before the diagnosis occurs, with symptoms occurring when a significant number of motor neurons are damaged.^24^ Conversely, cerebral ischemia caused by stroke may play a role in the development of ALS. Aggregation of misfolded proteins in neurons is part of the key pathophysiology of ALS, and ischemic stroke can induce the aggregation of ALS-related RNA-binding proteins such as TDP43 and FUS.^25^ Also, cerebral hypoxia induced by ischemic stroke leads to excessive reactive oxygen species production and neuronal cell death.^26^

The presence of physical disability in ALS patients did not affect the risk of ischemic stroke in the present study. The disabled ALS group, defined as patients with onset of physical disability within two years from the index date, suggests more rapid disease progression than the non-disabled ALS group. Previously, it was reported that a systemic low-grade inflammation in ALS patients, measured by the revised ALS Function Rating Scale (ALSFRS-R), was correlated with their degree of disability, suggesting that the systemic inflammatory state is apparently associated with a progression of and negative prognosis in ALS.^27^ In addition, disabled ALS patients can have more limited mobility, which is one of the risk factors for venous thromboembolism associated with ischemic stroke. The lack of an expected effect of disability on the risk of ischemic stroke in ALS patients may be due to the small number of events. Otherwise, this may suggest that inflammation and limited mobility, associated with disability, do not much contribute to the risk of ischemic stroke, although it needs to be further elucidated whether other possible mechanisms, such as TDP-43 proteinopathy or endothelial dysfunction, are enough to explain this relationship between ALS and ischemic stroke.

When stratified by age, sex, and BMI, the risk of ischemic stroke was higher in each subgroup of ALS patients than in the corresponding control group and was not different across all subgroups.

Our study has the following clinical implications. Careful history-taking and neurological examination are necessary for the early detection of ischemic stroke in ALS, since neurological changes by cerebral ischemia may be misdiagnosed as ALS progression. In addition, to prevent cerebral ischemia, prophylactic management of venous thromboembolism and appropriate treatment of acute ischemic stroke may be helpful in disabled ALS patients, but this theory needs to be further investigated. Moreover, the common mechanisms between ALS and ischemic stroke may help to reveal novel therapeutic targets based on inflammation, endothelial dysfunction, and TDP-43 proteinopathy.

This study also has several limitations. First, selection bias needs to be considered. Over 40% of both groups were excluded because they did not participate in national health check-up programs within two years from the index date. It is possible that patients who were more concerned about their health may have been included, and patients who had difficulty participating in health screenings due to severe physical disability may have been excluded. Second, administrative data were used; detailed clinical characteristics regarding the region of onset, disease duration, and medications for ALS treatment such as riluzole and edaravone, which can be neuroprotective in cerebral ischemia,^28, 29^ were not included in the analyses. Also, data regarding the subtype of ischemic stroke are absent. Therefore, subgroup analyses based on these factors could not be performed. Finally, physical disability in the NDRS is evaluated according to the Medical Research Council (MRC) scale and modified Barthel Index (mBI) as opposed to the ALSFRS-R, which is usually used to evaluate the function of ALS patients.^30^ With the limitations of the MRC scale and the mBI to evaluate respiratory and bulbar symptoms, bulbar-onset ALS patients may initially have mild limb weakness and could be classified into the non-disabled ALS group. However, limb-onset ALS accounts for two-thirds of all ALS cases,^1^ and the median time from onset to significant limb weakness in bulbar-onset ALS is approximately nine months.^31^ Furthermore, the MRC scale has been shown to be as good of a predictor of disease progression as forced vital capacity and ALSFRS-R, and the mBI is also strongly correlated with the ALSFRS-R.^32, 33^

In conclusion, we found that ALS patients face increased risk of ischemic stroke compared with controls, although the risk did not differ by the presence of disability in ALS patients. Further investigations are needed to elucidate the pathomechanism and potential risk factors of the increased risk of ischemic stroke in ALS.

## Data Availability

Data are available upon reasonable request. The data set analyzed in this study is not publicly available because of restricted access, but further information about the data set is available from the corresponding author on reasonable request.

## Contributors

Soonwook Kwon: Investigation, Writing – Original Draft. Bongseong Kim and Kyung-Do Han: Data Curation and Analysis. Wonyoung Jung and Eun Bin Cho: Investigation, Review. Dong Wook Shin and Ju-Hong Min (co-corresponding authors): Study Conception and Design, Validation, Supervision, Manuscript Correction.

## Acknowledgments

There are no acknowledgments.

## REFERENCES

1. Feldman EL, Goutman SA, Petri S, Mazzini L, Savelieff MG, Shaw PJ, et al. Amyotrophic lateral sclerosis. Lancet. 2022;400:1363–1380

2. Katsumata Y, Fardo DW, Kukull WA, Nelson PT. Dichotomous scoring of tdp-43 proteinopathy from specific brain regions in 27 academic research centers: Associations with alzheimer’s disease and cerebrovascular disease pathologies. Acta Neuropathol Commun. 2018;6:142

3. Huangfu N, Wang Y, Xu Z, Zheng W, Tao C, Li Z, et al. Tdp43 exacerbates atherosclerosis progression by promoting inflammation and lipid uptake of macrophages. Front Cell Dev Biol. 2021;9:687169

4. Pereira M, Gromicho M, Henriques A, Pronto-Laborinho AC, Grosskreutz J, Kuzma-Kozakiewicz M, et al. Cardiovascular comorbidities in amyotrophic lateral sclerosis. J Neurol Sci. 2021;421:117292

5. Diekmann K, Kuzma-Kozakiewicz M, Piotrkiewicz M, Gromicho M, Grosskreutz J, Andersen PM, et al. Impact of comorbidities and co-medication on disease onset and progression in a large german als patient group. J Neurol. 2020;267:2130–2141

6. Turner MR, Goldacre R, Talbot K, Goldacre MJ. Cerebrovascular injury as a risk factor for amyotrophic lateral sclerosis. J Neurol Neurosurg Psychiatry. 2016;87:244–246

7. Shin DW, Cho B, Guallar E. Korean national health insurance database. JAMA Intern Med. 2016;176:138

8. Cheol Seong S, Kim YY, Khang YH, Heon Park J, Kang HJ, Lee H, et al. Data resource profile: The national health information database of the national health insurance service in south korea. Int J Epidemiol. 2017;46:799–800

9. Jeong SM, Lee HR, Han K, Jeon KH, Kim D, Yoo JE, et al. Association of change in alcohol consumption with risk of ischemic stroke. Stroke. 2022;53:2488–2496

10. Kwon S, Jung SY, Han KD, Jung JH, Yeo Y, Cho EB, et al. Risk of parkinson’s disease in multiple sclerosis and neuromyelitis optica spectrum disorder: A nationwide cohort study in south korea. J Neurol Neurosurg Psychiatry. 2022

11. Kim M, Jung W, Kim SY, Park JH, Shin DW. The korea national disability registration system. Epidemiol Health. 2023:e2023053

12. Cho EB, Yeo Y, Jung JH, Jeong SM, Han K, Yang JH, et al. Acute myocardial infarction risk in multiple sclerosis and neuromyelitis optica spectrum disorder: A nationwide cohort study in south korea. Mult Scler. 2022;28:1849–1858

13. Jun KY, Park J, Oh KW, Kim EM, Bae JS, Kim I, et al. Epidemiology of als in korea using nationwide big data. J Neurol Neurosurg Psychiatry. 2019;90:395–403

14. Sundararajan V, Henderson T, Perry C, Muggivan A, Quan H, Ghali WA. New icd-10 version of the charlson comorbidity index predicted in-hospital mortality. J Clin Epidemiol. 2004;57:1288–1294

15. Sutedja NA, van der Schouw YT, Fischer K, Sizoo EM, Huisman MH, Veldink JH, et al. Beneficial vascular risk profile is associated with amyotrophic lateral sclerosis. J Neurol Neurosurg Psychiatry. 2011;82:638–642

16. Turner MR, Wotton C, Talbot K, Goldacre MJ. Cardiovascular fitness as a risk factor for amyotrophic lateral sclerosis: Indirect evidence from record linkage study. J Neurol Neurosurg Psychiatry. 2012;83:395–398

17. Gladman M, Dehaan M, Pinto H, Geerts W, Zinman L. Venous thromboembolism in amyotrophic lateral sclerosis: A prospective study. Neurology. 2014;82:1674–1677

18. Kupelian V, Viscidi E, Hall S, Li L, Eaton S, Dilley A, et al. Increased risk of venous thromboembolism in patients with amyotrophic lateral sclerosis: Results from a us insurance claims database study. Neurol Clin Pract. 2023;13:e200110

19. McCombe PA, Henderson RD. The role of immune and inflammatory mechanisms in als. Curr Mol Med. 2011;11:246–254

20. Chen S, Crother TR, Arditi M. Emerging role of il-17 in atherosclerosis. J Innate Immun. 2010;2:325–333

21. Lindsberg PJ, Grau AJ. Inflammation and infections as risk factors for ischemic stroke. Stroke. 2003;34:2518–2532

22. Garbuzova-Davis S, Woods RL, 3rd, Louis MK, Zesiewicz TA, Kuzmin-Nichols N, Sullivan KL, et al. Reduction of circulating endothelial cells in peripheral blood of als patients. PLoS One. 2010;5:e10614

23. Hainsworth AH, Oommen AT, Bridges LR. Endothelial cells and human cerebral small vessel disease. Brain Pathol. 2015;25:44–50

24. Eisen A, Kiernan M, Mitsumoto H, Swash M. Amyotrophic lateral sclerosis: A long preclinical period? J Neurol Neurosurg Psychiatry. 2014;85:1232–1238

25. Kahl A, Blanco I, Jackman K, Baskar J, Milaganur Mohan H, Rodney-Sandy R, et al. Cerebral ischemia induces the aggregation of proteins linked to neurodegenerative diseases. Sci Rep. 2018;8:2701

26. Mitroshina EV, Savyuk MO, Ponimaskin E, Vedunova MV. Hypoxia-inducible factor (hif) in ischemic stroke and neurodegenerative disease. Front Cell Dev Biol. 2021;9:703084

27. Keizman D, Rogowski O, Berliner S, Ish-Shalom M, Maimon N, Nefussy B, et al. Low-grade systemic inflammation in patients with amyotrophic lateral sclerosis. Acta Neurol Scand. 2009;119:383–389

28. Watanabe K, Tanaka M, Yuki S, Hirai M, Yamamoto Y. How is edaravone effective against acute ischemic stroke and amyotrophic lateral sclerosis? J Clin Biochem Nutr. 2018;62:20–38

29. Verma SK, Arora I, Javed K, Akhtar M, Samim M. Enhancement in the neuroprotective power of riluzole against cerebral ischemia using a brain targeted drug delivery vehicle. ACS Appl Mater Interfaces. 2016;8:19716–19723

30. Cedarbaum JM, Stambler N, Malta E, Fuller C, Hilt D, Thurmond B, et al. The alsfrs-r: A revised als functional rating scale that incorporates assessments of respiratory function. Bdnf als study group (phase iii). J Neurol Sci. 1999;169:13–21

31. Zhang H, Chen L, Tian J, Fan D. Disease duration of progression is helpful in identifying isolated bulbar palsy of amyotrophic lateral sclerosis. BMC Neurol. 2021;21:405

32. Magnus T, Beck M, Giess R, Puls I, Naumann M, Toyka KV. Disease progression in amyotrophic lateral sclerosis: Predictors of survival. Muscle Nerve. 2002;25:709–714

33. De Groot IJ, Post MW, Van Heuveln T, Van Den Berg LH, Lindeman E. Measurement of decline of functioning in persons with amyotrophic lateral sclerosis: Responsiveness and possible applications of the functional independence measure, barthel index, rehabilitation activities profile and frenchay activities index. Amyotroph Lateral Scler. 2006;7:167–172

